# Cathepsin G is associated with cerebral vascular injury in leukemia: A pathological insight into intracranial hemorrhage

**DOI:** 10.1101/2025.07.06.25330982

**Authors:** Toshihiro Gi, Kaiyou Kai, Kotaro Shide, Eriko Nakamura, Nobuyuki Oguri, Murasaki Aman, Kazunari Maekawa, Sayaka Moriguchi-Goto, Kazuya Shimoda, Yohei Hisada, Atsushi Yamashita

## Abstract

Intracranial hemorrhage (ICH) is a fatal complication of leukemia; however, the mechanisms underlying its development, particularly central nervous system (CNS) involvement and vascular injury, remain unclear. This autopsy-based study investigated the histopathological features of cerebral vessels in leukemia and the expression of hemostasis-related factors in leukemia cells. Thirty-eight leukemia cases and 20 matched controls were included. A histopathological analysis of CNS tissues was performed to evaluate ICH, leukemia cell localization, and vascular injury. Immunohistochemistry was conducted to assess the expression of vascular endothelial growth factor (VEGF), cathepsin G, tissue-plasminogen activator, urokinase-plasminogen activator, urokinase plasminogen activator receptor, and tissue factor in leukemia cells. Vascular integrity and permeability were evaluated using stains for smooth muscle actin, collagen, fibrin, and von Willebrand factor. ICH was identified in 66% of leukemia cases and was associated with fatal brain herniation in 40%. CNS involvement was observed in 59% of cases, often without a clinical diagnosis. The leukemia cell infiltration of meninges and vascular walls was frequently associated with changes in smooth muscle cells and adventitial collagen. CNS vascular injury was frequently associated with ICH in the presence of leukemia cell infiltration. VEGF and urokinase-plasminogen activator were highly expressed in leukemia cells. VEGF was associated with meningeal invasion, while cathepsin G was predominantly expressed in myeloid leukemia and linked to vascular damage. These results suggest that the leukemia cell infiltration of cerebral vascular walls plays an important role in cerebral vessel injury and leukemia cell-related ICH via VEGF and cathepsin G expression.

**Key points:**

- Leukemia cell infiltration into cerebral vascular walls is associated with vascular damage and intracranial hemorrhage in autopsy cases.
- The expression of VEGF and cathepsin G in leukemia may serve as markers of CNS involvement and cerebral vascular injury, respectively.

## Introduction

Intracranial hemorrhage (ICH) is a life-threatening complication in patients with hematological malignancies, particularly leukemia. In acute leukemia, the early mortality rate of patients who develop ICH was previously reported to be as high as 64%^1^, highlighting the importance of elucidating the mechanisms underlying this complication. Clinical studies investigated ICH in acute leukemia and identified a number of risk factors, including age, sex, the leukemia subtype, white blood cell count, platelet count, and prothrombin time.^2–6^

The majority of mechanistic studies have been conducted using leukemia murine models and peripheral blood from leukemia patients. These studies indicated the contribution of cytokines and hemostasis-related factors expressed by leukemia cells to coagulation abnormalities, the activation of fibrinolytic pathways, and vascular endothelial damage.^6–11^ These molecular abnormalities, together with thrombocytopenia caused by bone marrow suppression and an increased risk of infection, are considered to contribute to a complex bleeding diathesis. Therefore, ICH may predominantly be attributed to systemic bleeding tendency rather than a regional change in the central nervous system (CNS).

The local histopathology of hemorrhagic lesions in humans has not been examined in detail. Autopsy-based studies described ICH in leukemia patients and noted CNS involvement.^12–17^ The pathological findings of leukemia with ICH were identified as leukostasis, defined as dilated cerebral vessels densely packed with leukemia cells, and leukemic nodules, which are dense clusters of leukemia cells within the brain parenchyma.^12^ However, a detailed pathological evaluation of the affected vasculature and the expression of hemostatic factors in leukemia cells has not yet been performed. Therefore, the relationship between vascular infiltration and hemorrhage remains unclear. Hemostasis-related factors derived from leukemia cells may contribute to hemorrhagic complications. Vascular endothelial growth factor (VEGF), which increases vascular permeability, was shown to be expressed in leukemic blasts in both peripheral blood and bone marrow.^18^ Brain tumor-derived VEGF can compromise vascular function and promote hemorrhage in a murine model.^19^ Furthermore, cathepsin G, a serine protease highly expressed in leukemia, induced endothelial injury in *in vitro* cell models.^20,21^ Elevated levels of fibrinolysis-related factors have also been detected in the peripheral blood of patients with acute leukemia, and have been associated with bleeding tendency.^22^ Based on these findings, we hypothesized that the expression of these factors in leukemia cells may contribute to the development of ICH.

Therefore, the present study investigated histopathological changes in cerebral vessels in human autopsy specimens to clarify the expression of hemostasis-related factors in leukemia cells, the direct involvement of leukemia cells in ICH, and their potential impact on vascular integrity.

## Materials and Methods

### Selection of leukemia cases

This retrospective study was approved by the Ethics Committee of the University of Miyazaki (protocol number: O-1417). The study design is summarized in Figure S1. We selected leukemia cases (n=54) from consecutive autopsy cases at the University of Miyazaki Hospital between 1977 and 2023 (n=2,673). Sixteen cases without a CNS examination were excluded. We also selected autopsy cases without CNS diseases for the control group (n=20), matched by age and sex with the leukemia group. Cases in the control group had no history of hematological malignancies or CNS diseases, such as stroke, cancer metastasis to the CNS, neurodegenerative diseases, or CNS infections. We analyzed the clinicopathological backgrounds, histological types of leukemia, and presence of ICH based on autopsy records. We then performed a histopathological analysis of available hematoxylin and eosin (HE)-stained brain specimens. We also conducted immunohistochemistry and immunofluorescence using paraffin-embedded tissues of the brain and systemic organs.

### Histological analysis with HE-stained sections

All specimens were fixed in 20–30% formaldehyde, embedded in paraffin, sectioned at a thickness of 2.5 μm, and stained with HE. The presence and distribution of leukemia cells in brain tissue were analyzed. The anatomical sites of leukemia cell localization were classified as follows: the subarachnoid space extending to the Virchow-Robin space, the intravascular space, and the brain parenchyma (Figure 1A). CNS involvement was defined as the presence of leukemia cells in any of these sites. Infiltration within the subarachnoid space extending to the Virchow–Robin space was regarded as meningeal invasion by leukemia. We also investigated whether pathological leukostasis and leukemic nodules were present, as previously described.^12^ Briefly, pathological leukostasis was defined as dilated cerebral vessels densely packed with leukemia cells, and leukemic nodules as dense clusters of leukemia cells within the brain parenchyma.^12^ The sites of ICH were classified as follows: the subarachnoid space extending to the Virchow-Robin space, the brain parenchyma, the intraventricular space, and the subdural region. The causes of ICH were examined using autopsy records and assessments of tissue sections, and classified into the following categories: systemic bleeding tendency, CNS involvement of leukemia, aspergillosis, and hemorrhagic infarction. ICH caused by CNS involvement was defined as the presence of dense leukemia cell infiltration in cerebral vessels at hemorrhagic sites with or without systemic bleeding tendency.

**Figure 1.**
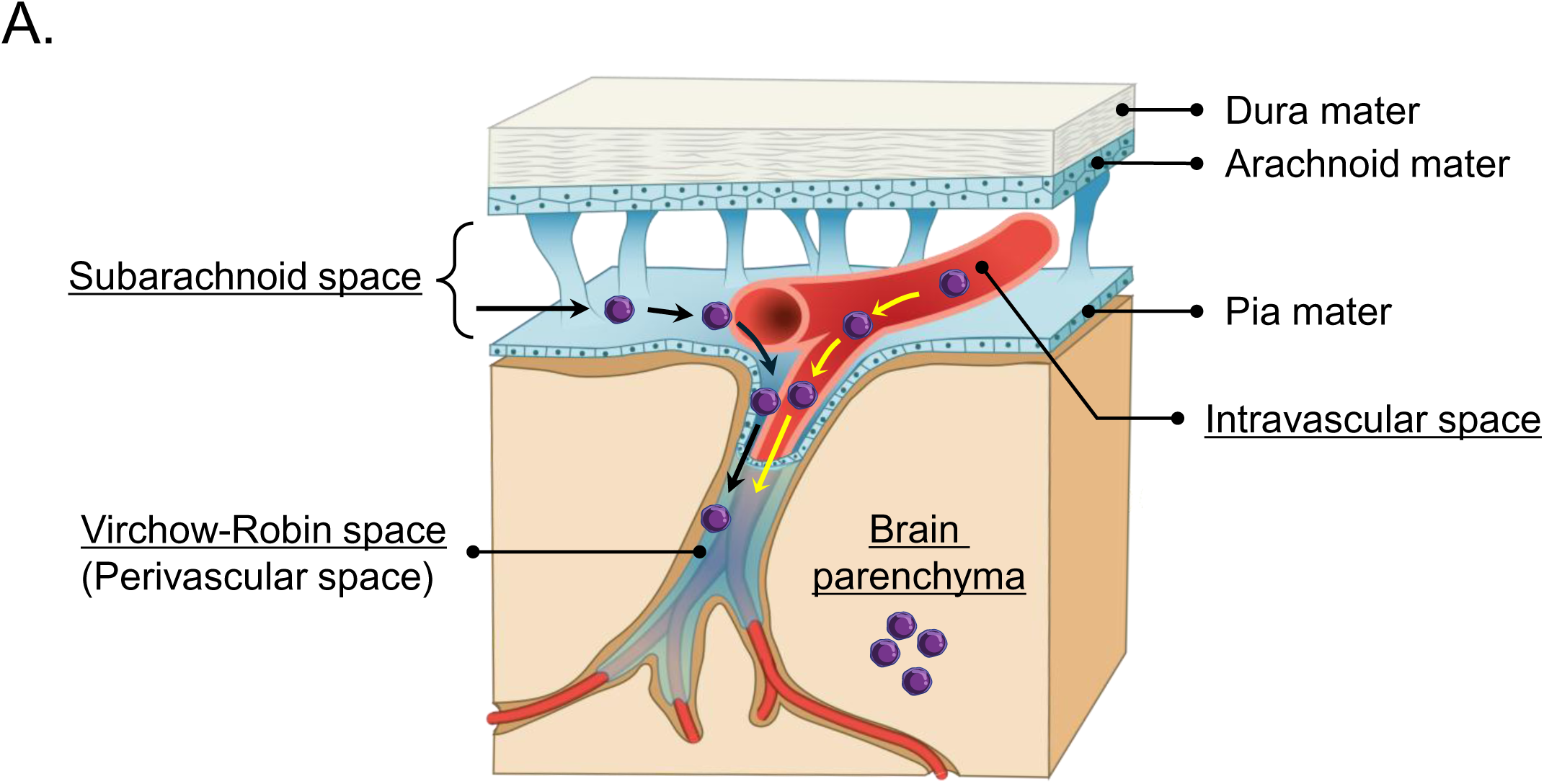
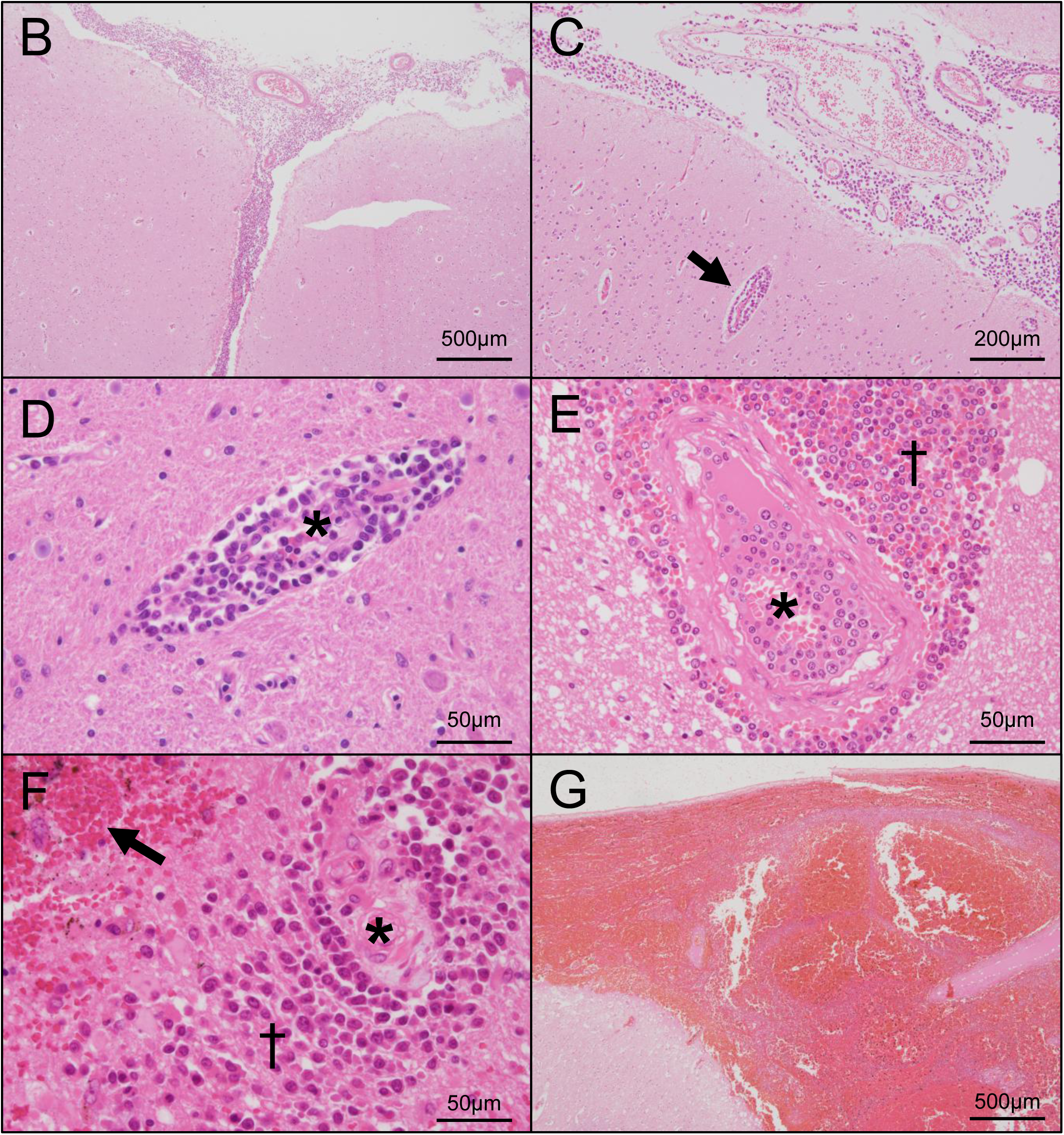
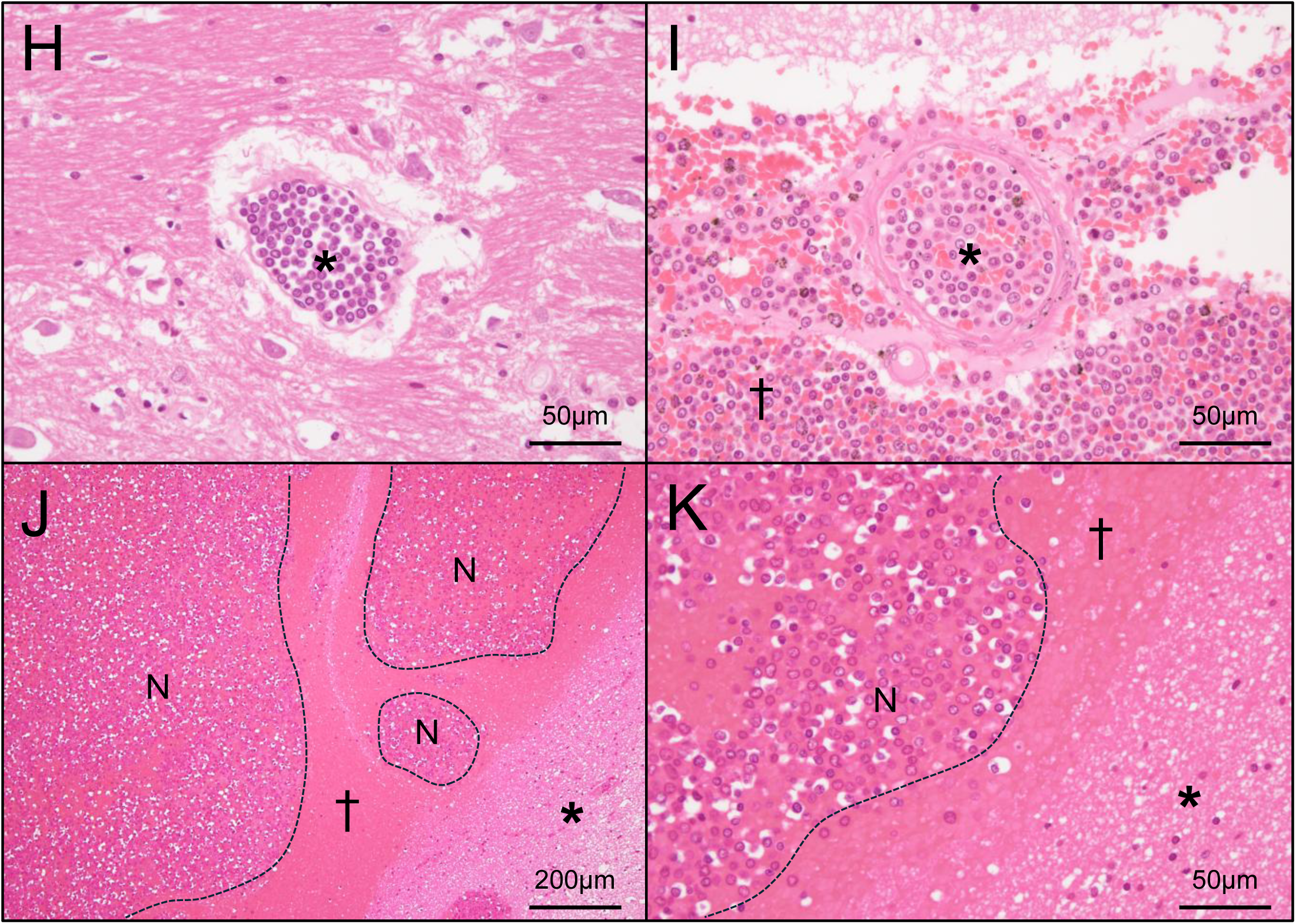
Patterns of CNS involvement in leukemia and intracranial hemorrhage. **A.** Schematic representation of the meningeal structure and brain vessel distribution. Leukemia cells infiltrate the central nervous system via cerebrospinal fluid or blood flow. Leukemia cells disseminated through cerebrospinal fluid flow extend into the subarachnoid space and Virchow-Robin space. In this study, the localization of leukemia cells was classified as follows: 1) the subarachnoid space extending into the Virchow-Robin space, 2) the intravascular space, and 3) the brain parenchyma. **B–D.** Pathological patterns of CNS involvement in leukemia. **B.** Leukemia cell infiltration in the subarachnoid space in a case of ALL, consistent with meningeal invasion. **C.** Leukemia cell infiltration along the subarachnoid space and Virchow-Robin space (VRS; arrow) in a case of AML. **D.** Leukemia cell infiltration in the VRS in a case of AML. Leukemia cells are localized in the VRS without brain parenchymal involvement. The asterisks indicate the vascular lumens (D and E). **E–G.** Pathological patterns of intracranial hemorrhage. **E.** Subarachnoid hemorrhage in a case of CML-BP with fatal brain herniation. Hemorrhage extends into the VRS (†), accompanied by dense leukemia cell clusters. Leukemia cells are also present in the intravascular space (asterisk). **F.** Brain parenchymal hemorrhage in a case of AML. A vessel is indicated by an asterisk. Presence of leukemia cells in the VRS and parenchyma (†), accompanied by hemorrhage (arrow). **G.** Subarachnoid hemorrhage in a case of AML with fatal brain herniation. **H and I.** Pathological leukostasis in brain vessels in cases of AML (H) and CML in the blast phase (I). Intravascular spaces are filled with leukemia cells, with few or no erythrocytes visible (H and I, asterisks). In panel I, a vessel with leukostasis is observed within a subarachnoid hemorrhagic site infiltrated by leukemia cells (†). **J and K.** Leukemic nodules in intracranial hemorrhagic sites in a case of AML. In the low-magnification field (J), multinodular, hypercellular lesions (N, within dashed lines) are observed within a hemorrhagic area (†). In the high-magnification field (K), these nodular lesions (N) consist of densely packed leukemia cells. Asterisks indicate glial tissue (J and K).

### Immunohistochemical examination of leukemia cells and cerebral vessels

We focused on the expression of hemostatic factors in leukemia cells. The primary antibodies used are described in Table S1. Representative paraffin sections containing leukemia cells from each case were stained with VEGF (rabbit polyclonal, A-20; Santa Cruz Biotechnology), cathepsin G (rabbit, monoclonal, clone E3N30; Cell Signaling Technology, MA, USA), tissue-plasminogen activator (tPA; rabbit polyclonal; GeneTex, Irvine, California, USA), urokinase-plasminogen activator (uPA; rabbit polyclonal; Atlas Antibodies, Stockholm, Sweden), urokinase-plasminogen activator receptor (uPAR; mouse, monoclonal, clone E-3; Santa Cruz Biotechnology), and tissue factor (TF; mouse monoclonal, clone H-9; Santa Cruz Biotechnology, Dallas, TX, USA). In cases without leukemia cells in the brain, representative paraffin blocks from other organs containing leukemia cells were selected. We defined a sample as immunopositive when more than 10% of leukemia cells were stained for a given protein, including VEGF, cathepsin G, tPA, uPA, uPAR, or TF, as previously report.^23^ Representative AML cases were also stained by myeloperoxidase (MPO, rabbit, polyclonal, Dako/Agilent, Santa Clara, CA, USA). We then performed Sirius Red staining and immunohistochemistry to examine vascular integrity and permeability. Brain sections from the leukemia and control groups were stained for α-smooth muscle actin (SMA; mouse monoclonal antibody, clone 1A4; Dako/Agilent, Santa Clara, CA, USA), fibrin (mouse monoclonal antibody, clone 59D8; EMD Millipore, Burlington, MA, USA), and von-Willebrand factor (VWF; mouse monoclonal, clone 36B11; Leica Biosystems, IL, USA). Sirius Red staining and immunohistochemistry for SMA were performed to assess vascular collagen fibers and smooth muscle cells, respectively. Collagen fibers were identified as regions exhibiting an orange-to-green color shift under a polarized lens. Abnormal vascular permeability was defined by the presence of fibrin or VWF deposition within the vascular wall or in the perivascular region. Sections were stained with EnVision, a dextran polymer conjugated with anti-mouse or rabbit immunoglobulin and horseradish peroxidase (DAKO/Agilent). Horseradish peroxidase activity was visualized using 3,3’-diaminobenzidine solution containing hydrogen peroxide, and sections were counterstained with Mayer’s hematoxylin. Control sections were stained with non-immune mouse or rabbit immunoglobulin.

We performed double immunofluorescence to examine cathepsin G and VEGF expression in leukemia cells in representative brain specimens. Brain sections were stained with MPO light chain (mouse monoclonal, clone A-5; Santa Cruz Biotechnology) and cathepsin G (clone E3N30) or VEGF (A-20). CF568 conjugated-donkey anti-mouse IgG (Biotium, Hayward, CA, USA) and CF488 conjugated-donkey anti-rabbit IgG (Biotium) were used as secondary antibodies. Brain sections were mounted using 4, 6’-diamidino-2-phenylindole-containing reagents. Fluorescent images were captured using an all-in-one fluorescence microscope (BZ-X810; Keyence, Osaka, Japan). Image acquisition and channel merging were performed using the microscope’s dedicated software (BZ-X Series Application, Keyence).

### Statistical analysis

Categorical data were analyzed using Fisher’s exact test. Data are presented as medians and ranges. Since all data showed a non-normal distribution, the Mann-Whitney U test was used for comparisons of two groups. Statistical analyses were performed using GraphPad Prism 8.43 (GraphPad Software Inc., San Diego, CA, USA). A p-value < 0.05 was considered to be significant.

## Results

### Clinicopathological characteristics of leukemia cases

Table 1 summarizes the clinicopathological characteristics of leukemia cases in autopsy records (n=38). Median age was 58 years, with a male-to-female ratio of 3:1. A history of chemotherapy for leukemia was noted in 87% of cases. Leukemia subtypes included acute myeloid leukemia (AML; n=25, 66%), acute lymphoblastic leukemia (ALL; n=7, 18%), chronic myeloid leukemia in the blast phase (CML-BP; n=5, 13%), and chronic lymphocytic leukemia/small lymphocytic lymphoma (CLL/SLL; n=1, 3%). The CNS involvement of leukemia cells was clinically diagnosed in 8% (3/38) of cases. Specifically, the diagnoses were based on the following findings in each individual case. The first case: seizures, electroencephalography, and a cerebrospinal fluid examination; The second case: seizure episodes alone; and the third case: a cerebrospinal fluid examination and magnetic resonance imaging. Hyperleukocytosis at death (white blood cell count >100,000/µL) was observed in 13% (5/38) of cases. A post-mortem examination revealed that 90% of cases (n=34) were in a non-remission state. Pathological CNS involvement was noted in 53% (20/38). ICH and cerebral infarction were identified in 66% (25/38) and 8% (3/38) of cases, respectively. Severe infection was present in 63% (24/38), with fungal infections occurring in 26% (10/38). Systemic hemorrhage and multiple microthrombi were detected in 53% (20/38) and 13% (5/38) of cases, respectively. No cases of deep vein thrombosis were identified. Pulmonary embolism was found in four cases (11%), all of which were caused by fungal emboli. Non-bacterial thrombotic endocarditis (NBTE) was observed in 5% (2/38), with one case complicated by fatal hemorrhagic infarction. Fatal brain herniation due to ICH was identified in 24% (9/38) of cases. Table S2 shows the clinicopathological characteristics of the control group. There were no significant differences in age, sex, severe infection, systemic hemorrhage, pulmonary embolism, or NBTE between the control and leukemia groups (Table S3). A history of chemotherapy was more frequent in the leukemia group than in the control group (Table S3). DVT was more frequent in the control group (Table S3).

**Table 1.** Clinicopathological findings of autopsy cases of leukemia (n=38)

| Clinical background |  |  | Autopsy findings, n (%) |  |
| --- | --- | --- | --- | --- |
| Age, y, median (range) | 58 | (13-82) | Non-remission state of leukemia | 34 (89.5) |
| Male, n (%) | 29 | (76.3) | *CNS involvement of leukemia cells* | 20 (52.6) |
| Chemotherapy, n (%) | 33 | (86.8) | Intracranial hemorrhage | 25 (65.8) |
| Leukemia type, n (%) |  |  | Cerebral infarction | 3 (7.9) |
| AML | 25 | (65.8) | Severe infection‡ | 24 (63.2) |
| M2 | 7 | (18.4) | Fungal infection | 10 (26.3) |
| M3/APL | 2 | (5.3) | *Aspergillus* | 6 (15.8) |
| M5 | 5 | (13.2) | *Aspergillus and Mucor* | 2 (5.3) |
| Unclassified/Unknown | 11 | (28.9) | *Candida* | 1 (2.6) |
| ALL | 7 | (18.4) | *Candida and Mucor* | 1 (2.6) |
| CML, blast phase | 5 | (13.2) | Systemic hemorrhage | 20 (52.6) |
| CLL/SLL | 1 | (2.6) | Multiple microthrombi§ | 5 (13.2) |
| Clinical diagnosis of CNS involvement | 3 | (7.9) | DVT | 0 (0) |
| Hyperleukocytosis at death* | 5 | (13.2) | PE\|\| | 4 (10.5) |
|  |  |  | NBTE | 2 (5.3) |
|  |  |  | Fatal brain herniation | 10 (26.3) |
|  |  |  | *Caused by intracranial hemorrhage* | 9 (23.7) |
|  |  |  | *Caused by hemorrhagic infarction with NBTE* | 1 (2.6) |

DVT, deep vein thrombosis; PE, pulmonary embolism; NBTE, non-bacterial thrombotic endocarditis. \*Hyperleukocytosis was defined as a white blood cell count >100,000/ μL. †Severe infection was defined as an infectious disease diagnosed clinically and pathologically, which was related to the cause of death.
‡Multiple microthrombi was defined as the presence of microthrombi in more than three organs. §All PE cases were caused by fungal emboli.

### Pathological findings in leukemia complicated by ICH

Table S4 shows the clinicopathological findings of leukemia cases complicated by ICH (n=25). Among leukemia types, ICH was frequently observed in AML (n=16, 64%). A non-remission state was noted in 92% (23/25) of cases, with the CNS involvement of leukemia in 44% (11/25). The locations of ICH included the brain parenchyma (76%, 19/25), the subarachnoid space extending to the Virchow-Robin space (64%, 16/25), the intraventricular space (28%, 7/25), the subdural region (12%, 3/25), and multiple sites (60%, 15/25). Fatal brain herniation occurred in 40% (10/25) of cases. The causes of ICH were as follows: systemic bleeding tendency (52%, 13/25), the CNS involvement of leukemia (32%, 8/25), aspergillosis (8%, 2/25), emboli from NBTE (4%, 1/25), and the co-occurrence of mucormycosis and leukemia involvement (4%, 1/25).

### Pathological findings in leukemia with CNS involvement

Table 2 shows the characteristics of leukemia cases with pathological CNS involvement (n=20). Only 5% (1/20) of cases were clinically diagnosed with CNS involvement. Hyperleukocytosis at death was observed in 20% (4/20) of cases. CNS involvement was most frequently observed in AML cases. Leukemia cells were detected in the subarachnoid space extending to the Virchow-Robin space (95%, 19/20; Figures 1B-1D), the intravascular space (50%, 10/20; Figure 1E), and the brain parenchyma (35%, 7/20, Figure 1F). A single case (5%, 1/20) had leukemia cells in the intravascular space only. ICH was noted in 50% (10/20) of cases with CNS involvement and was accompanied by leukemia cell infiltration at hemorrhagic sites in 40% (8/20) of cases (Figures 1E-1G). Leukostasis and leukemic nodules were each observed in 20% (4/20) of cases (Figures 1H-1K). Three cases exhibited both leukostasis and leukemic nodules, while one case showed leukostasis alone and another showed only leukemic nodules. All leukemic nodules were located within hemorrhagic areas (Figures 1J and 1K). Leukostasis in two cases and leukemic nodules in one case were accompanied by hyperleukocytosis.

**Table 2.** Characteristics of leukemia cases with pathological CNS involvement (n=20), n (%)

|  |  |  |
| --- | --- | --- |
| Clinical diagnosis of CNS involvement | 1 | (5.0) |
| Hyperleukocytosis at death* | 4 | (20.0) |
| Leukemia type |  |  |
| AML | 12 | (60.0) |
| ALL | 5 | (25.0) |
| CML, blast phase | 3 | (15.0) |
| Sites of leukemia cells in CNS† |  |  |
| Subarachnoid extending to the Virchow-Robin space | 19 | (95.0) |
| Intravascular space | 10 | (50.0) |
| Brain parenchyma | 7 | (35.0) |
| Intracranial hemorrhage | 10 | (50.0) |
| *accompanied by leukemia cell infiltration* | 8 | (40.0) |
| *in hemorrhagic sites* |  |  |
| *complicated with fetal brain herniation* | 4 | (20.0) |
| Intracranial leukostasis | 4 | (20.0) |
| *complicated with hyperleukocytosis at death* | 2 | (10.0) |
| Leukemic nodules | 4 | (20.0) |
| *complicated with hyperleukocytosis at death* | 1 | (5.0) |

\*Hyperleukocytosis was defined as a white blood cell count >100,000/ μL.
†Cases involved multiple sites

### Expression of hemostasis-related factors in leukemia cells

We examined the immunohistochemical expression of VEGF, cathepsin G, tPA, uPA, uPAR, and TF in leukemia cells in 28 cases with leukemia specimens. Immunohistochemistry was performed on brain specimens with leukemia cells or on representative specimens from other affected organs (lymph node, spleen, bone marrow, kidney, or lung) if brain involvement was absent. Table S5 summarizes the frequency of immunohistochemical expression in leukemia cells according to CNS involvement and the leukemia type. VEGF, cathepsin G, tPA, uPA, uPAR, and TF expression was observed in 82, 32, 4, 79, 11, and 7% of cases, respectively (Table S5; Figures 2A and 2B). Among the leukemia types, VEGF and uPA were frequently expressed in all types, except for CLL/SLL. The expression of cathepsin G, a neutrophil serine protease, was observed in 41% of AML cases and in 50% of CML-BP cases, but not in ALL or CLL/SLL cases. In double immunofluorescence, MPO-positive myeloid leukemia cells heterogeneously expressed VEGF or cathepsin G (Figures 2C and 2D). TF was rarely expressed or absent in all leukemia types.

**Figure 2.**
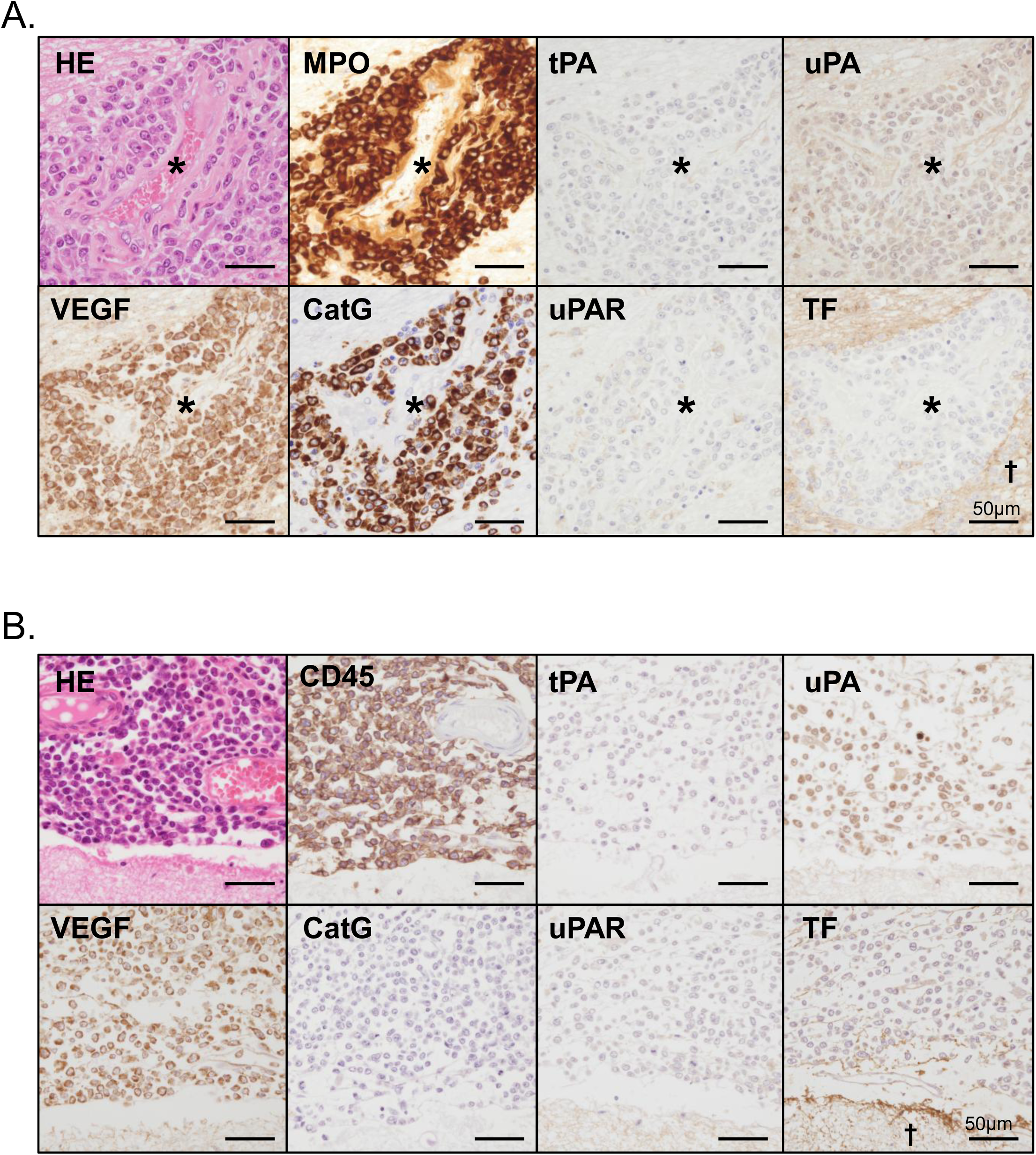
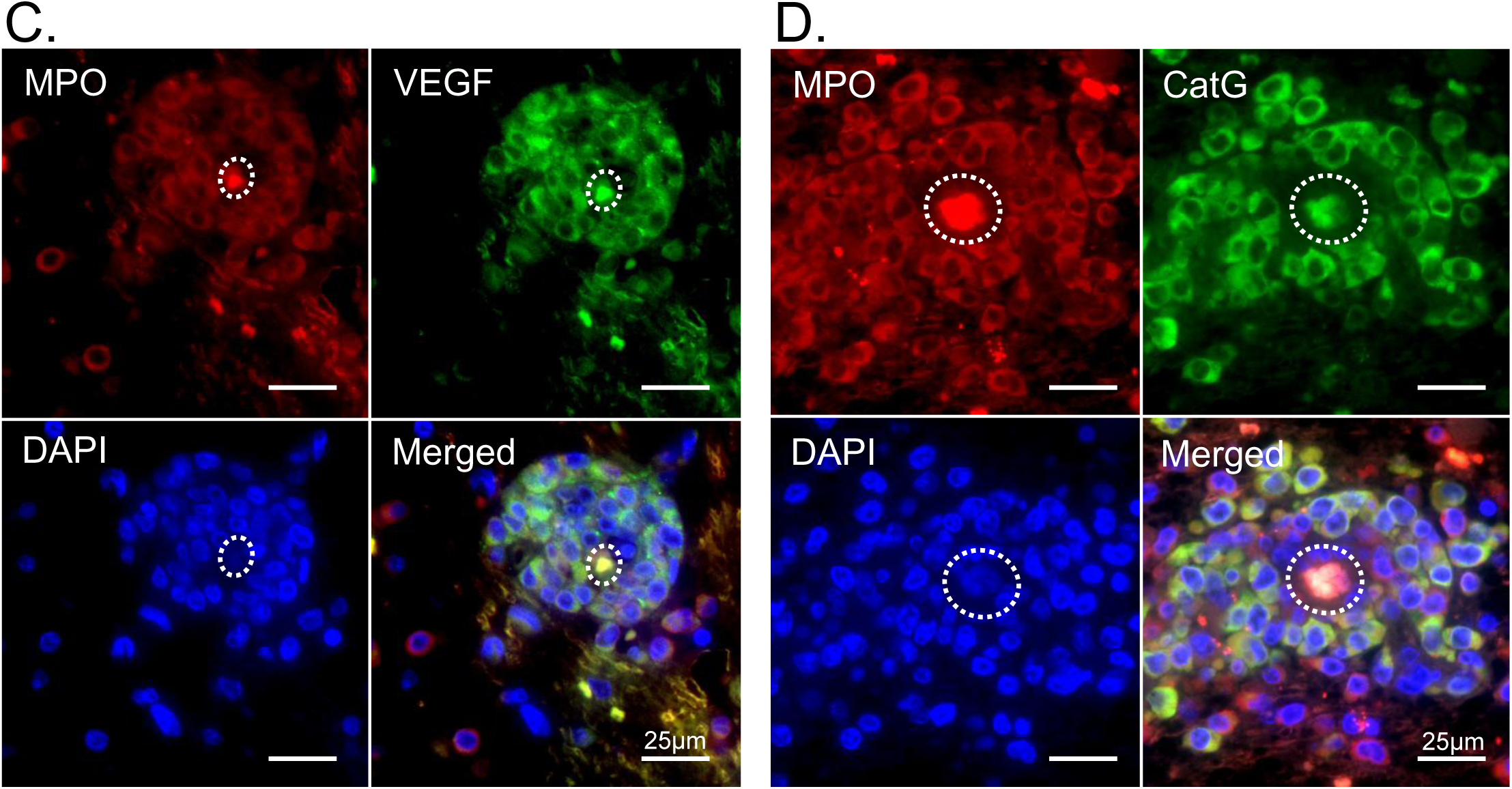
Expression of hemostatic factors in leukemia cells. **A.** Leukemia cell infiltration in the Virchow-Robin space in a case of AML. Asterisks indicate the vascular lumen. Myeloperoxidase (MPO)-positive leukemia cells express vascular endothelial growth factor (VEGF) and cathepsin G (CatG), but are negative for tissue plasminogen activator (tPA), urokinase-type plasminogen activator (uPA), urokinase-type plasminogen activator receptor (uPAR), and tissue factor (TF). **B.** Leukemia cell infiltration in the subarachnoid space in a case of ALL. CD45-positive leukemia cells express VEGF and uPA, but are negative for CatG, tPA, uPA, uPAR, and TF. TF expression is observed in the surrounding glial tissue (†). **C and D.** Double immunofluorescence of leukemia cells in the brain (same case as in Figure 2A). MPO-positive leukemia cells show the heterogeneous expression of VEGF (C) and CatG (D). Dashed lines indicate the vascular lumen. DAPI, 4′,6-diamidino-2-phenylindole.

### Vascular injury and abnormal permeability of cerebral vessels

Since leukemia cells were characteristically observed along and within cerebral vessels, we examined vascular injury and the abnormal permeability of cerebral vessels in the leukemia group (n=38) and control group (n=20). Vascular injury was defined as the disruption or fragmentation of SMA-positive or Sirius Red-positive layers. Abnormal vascular permeability was defined as fibrin or VWF deposition in intravascular or perivascular regions. The medial smooth muscle cell layer and adventitial collagen layer were preserved in the control group. Furthermore, vascular injury was not observed in the control group (0/20; Figure 3A). In contrast, vascular injury was detected in 34% (13/38) of leukemia cases. All cases with vascular injury were associated with the direct infiltration of leukemia cells into the vascular walls, either in the brain parenchyma or subarachnoid space (Figures 3B and 3D). No abnormal vascular permeability was noted in the control group (0/20; Figure 3C). Fibrin or VWF deposition in intravascular or perivascular regions was observed in 16% (6/38) of leukemia cases (Figure 3D).

**Figure 3.**
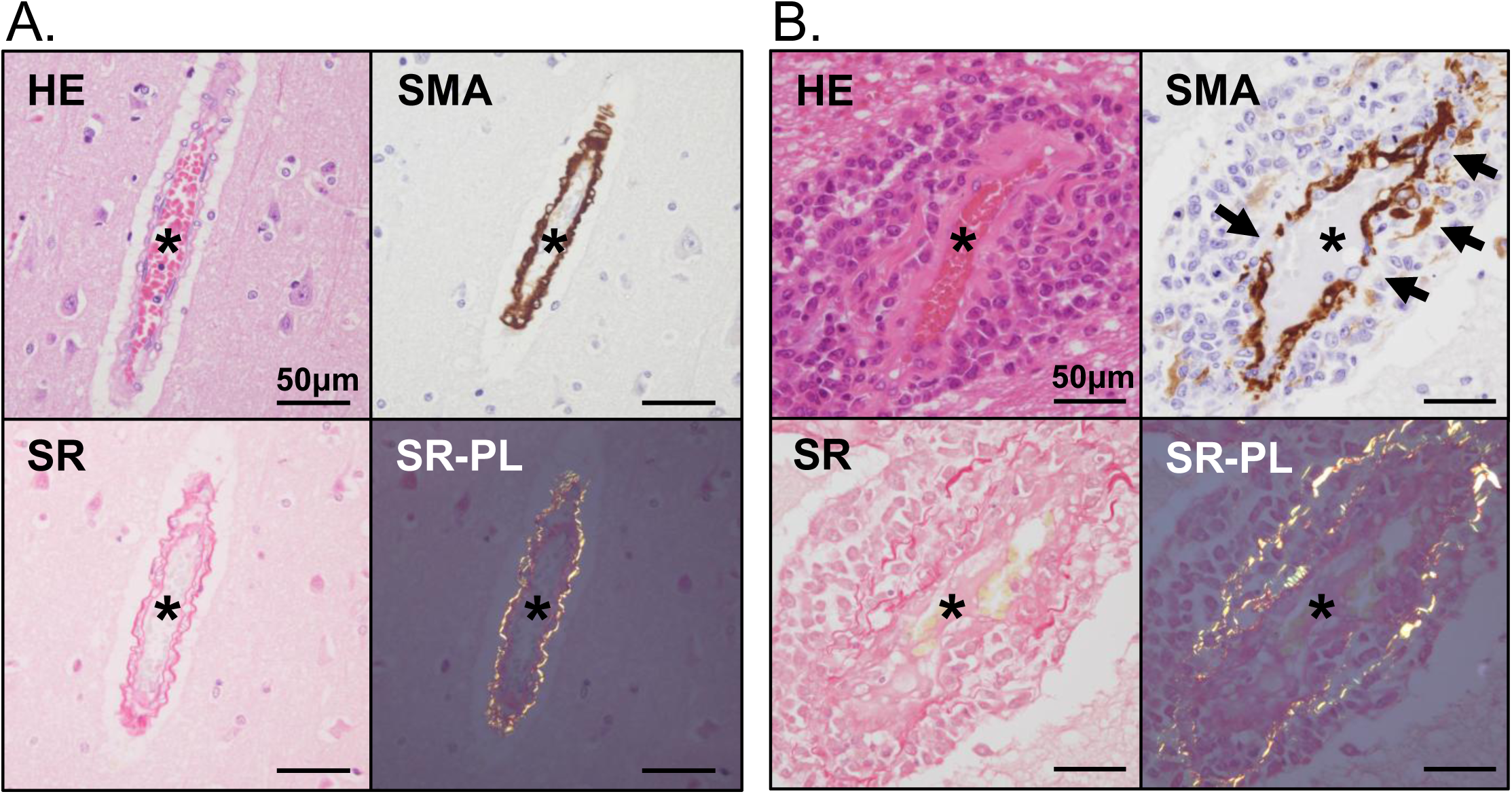
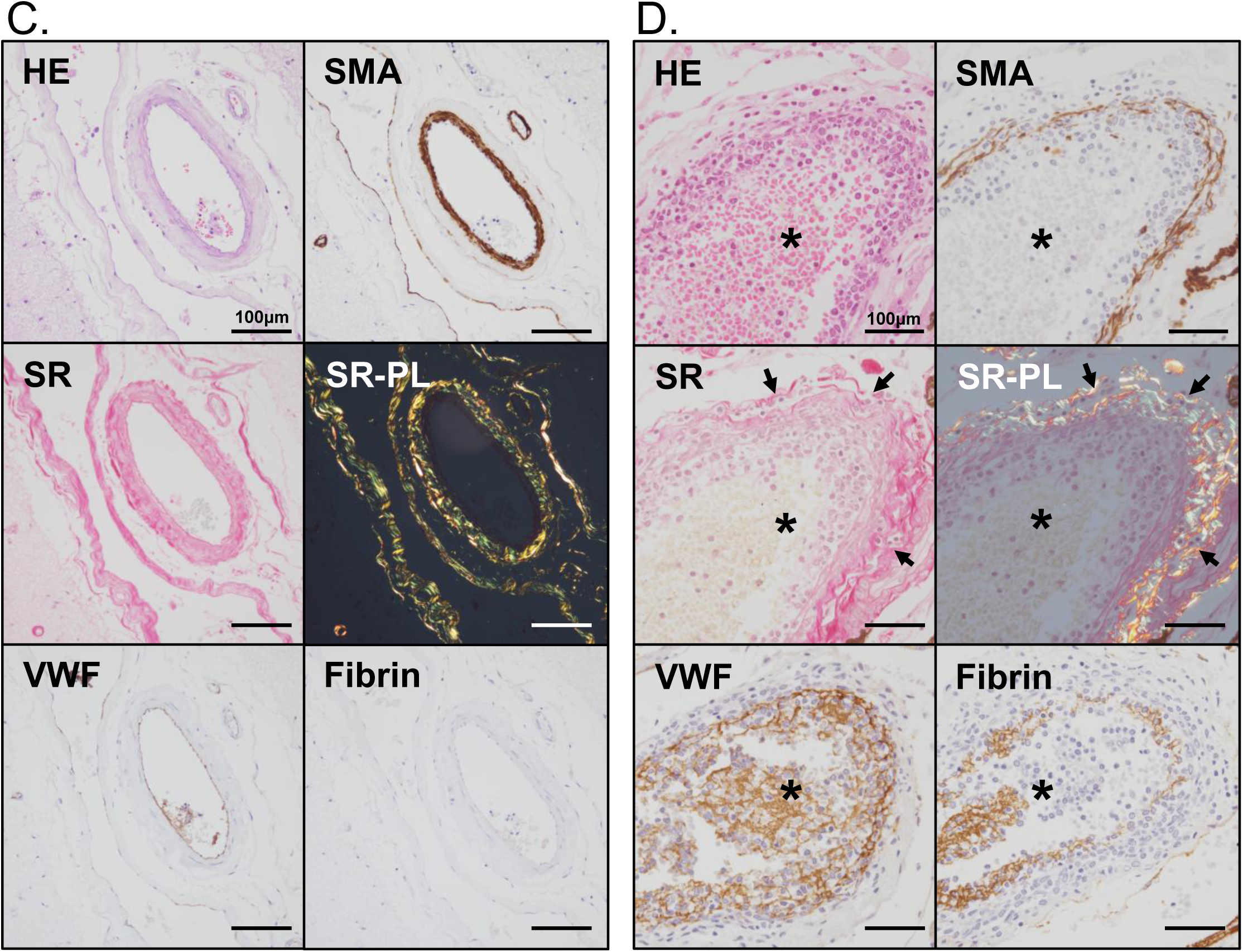
Direct vascular injury and abnormal permeability. **A.** Histology of a cerebral vessel in the control brain. Smooth muscle layers and collagen fibers are highlighted by SMA and SR, respectively. In contrast to panel A, no disruption or fragmentation of SMA-positive smooth muscle cells or collagen fibers is observed. Additionally, no cellular infiltration is noted in the Virchow–Robin space. **B.** Leukemia cell infiltration in the Virchow-Robin space in a case of AML. This is the same case as shown in Figure 2A. The smooth muscle layer of the vessel wall is immunopositive for smooth muscle actin (SMA). SMA-immunopositive cells exhibit disruption and fragmentation, associated with leukemia cell involvement (arrows). Similarly, collagen fibers in the vessel wall, highlighted by Sirius Red staining (SR), appear fragmented due to leukemia cell infiltration. Asterisks indicate the vascular lumen (A and B). SR-PL, Sirius Red staining observed under a polarized lens. **C.** Histology of a cerebral vessel in the subarachnoid space of a control brain. In contrast to panel C, no disruption or fragmentation of SMA-positive smooth muscle cells or collagen fibers is observed. VWF is expressed in endothelial cells, with no abnormal deposition of VWF or fibrin within the vascular wall or surrounding tissue. **D.** Leukemia cell infiltration into a vessel in the subarachnoid space in a case of AML. Leukemia cells infiltrate between SMA-positive fibers, leading to their fragmentation. Some leukemia cells are also observed between collagen fibers, highlighted by SR (arrows). In this case, leukemia cells are predominantly located on the luminal side. VWF and fibrin are widely distributed within the vascular wall. Immunopositive staining for VWF and fibrin is also observed in the lumen (asterisks), indicating small thrombus formation. SR-PL, Sirius Red staining observed under a polarized lens.

### Relationships among ICH, CNS involvement, and expression of hemostatic factors

We examined the relationships among ICH, CNS involvement, and the expression of hemostatic factors. No significant differences were noted in the frequency of CNS involvement, vascular injury, or abnormal permeability between the non-ICH and ICH groups (Table S6). Fibrinolysis- and vascular permeability-related factors, particularly uPA and VEGF, were frequently expressed, regardless of the ICH status (Table S6). All cases exhibiting pathological leukostasis or leukemic nodules were complicated by ICH (Table S6); however, no significant differences were noted between the ICH and non-ICH groups.

Among leukemia cases with CNS involvement (n=20), cathepsin G expression was more frequent in cases with vascular injury than in those without vascular injury (54% vs. 0%, p=0.045; Table 3). VEGF expression was significantly more frequent in cases with meningeal invasion than in those without meningeal invasion (95% vs. 56%, p=0.026; Table 3). The expression of other hemostatic factors did not significantly differ in cases with or without vascular injury, abnormal permeability, CNS involvement, or meningeal invasion (Table 3).

**Table 3.** Relationships among vascular injury, CNS involvement, and the expression of hemostatic factors, n (%)

| Leukemia-expressing factors in the brain* | Total (n=20) |  | Non-vascular injury |  | Vascular injury |  | p value |
| --- | --- | --- | --- | --- | --- | --- | --- |
|  |  |  | (n=7) |  | (n=13) |  |  |
| TF | 0 | (0) | 0 | (0) | 0 | (0) | 1 |
| tPA | 1 | (5.0) | 1 | (14.3) | 0 | (0) | 0.35 |
| uPA | 16 | (80.0) | 6 | (85.7) | 10 | (76.9) | 1 |
| uPAR | 1 | (5.0) | 0 | (0) | 1 | (7.7) | 1 |
| Cathepsin G | 7 | (35.0) | 0 | (0) | 7 | (53.8) | **0.045** |
| VEGF | 18 | (90.0) | 6 | (85.7) | 12 | (92.3) | 1 |

| Leukemia-expressing factors in the brain* | Total (n=20) |  | Normal permeability |  | Abnormal permeability (n=4) |  | p value |
| --- | --- | --- | --- | --- | --- | --- | --- |
|  |  |  | (n=16) |  |  |  |  |
| TF | 0 | (0) | 0 | (0.0) | 0 | (0) | 1 |
| tPA | 1 | (5.0) | 1 | (6.3) | 0 | (0) | 1 |
| uPA | 16 | (80.0) | 13 | (81.3) | 3 | (75.0) | 1 |
| uPAR | 1 | (5.0) | 0 | (0) | 1 | (25.0) | 0.20 |
| Cathepsin G | 7 | (35.0) | 5 | (31.3) | 2 | (50.0) | 0.59 |
| VEGF | 18 | (90.0) | 14 | (87.5) | 4 | (100.0) | 1 |

| Leukemia-expressing factors in any organ† | Total (n=28) |  | Normal permeability |  | Abnormal permeability (n=4) |  | p value |
| --- | --- | --- | --- | --- | --- | --- | --- |
|  |  |  | (n=24) |  |  |  |  |
| TF | 2 | (7.1) | 2 | (8.3) | 0 | (0) | 1 |
| tPA | 1 | (3.6) | 1 | (4.2) | 0 | (0) | 1 |
| uPA | 22 | (78.6) | 19 | (79.2) | 3 | (75.0) | 1 |
| uPAR | 3 | (10.7) | 2 | (8.3) | 1 | (25.0) | 0.38 |
| Cathepsin G | 9 | (32.1) | 7 | (29.2) | 2 | (50.0) | 0.57 |
| VEGF | 23 | (82.1) | 19 | (79.2) | 4 | (100.0) | 1 |

| Leukemia-expressing factors in any organ† | Total (n=28) |  | Non-CNS involvement (n=8) |  | CNS involvement (n=20) |  | p value |
| --- | --- | --- | --- | --- | --- | --- | --- |
| TF | 2 | (7.1) | 2 | (25.0) | 0 | (0) | 0.074 |
| tPA | 1 | (3.6) | 0 | (0) | 1 | (5.0) | 1 |
| uPA | 22 | (78.6) | 6 | (75.0) | 16 | (80.0) | 1 |
| uPAR | 3 | (10.7) | 2 | (25.0) | 1 | (5.0) | 0.19 |
| Cathepsin G | 9 | (32.1) | 2 | (25.0) | 7 | (35.0) | 1 |
| VEGF | 23 | (82.1) | 5 | (62.5) | 18 | (90.0) | 0.12 |

| Leukemia-expressing factors in any organ† | Total (n=28) |  | Non-meningeal invasion (n=9) |  | Meningeal invasion (n=19) |  | p value |
| --- | --- | --- | --- | --- | --- | --- | --- |
| TF | 2 | (7.1) | 2 | (22.2) | 0 | (0) | 0.10 |
| tPA | 1 | (3.6) | 0 | (0) | 1 | (5.3) | 1 |
| uPA | 22 | (78.6) | 7 | (77.8) | 15 | (78.9) | 1 |
| uPAR | 3 | (10.7) | 2 | (22.2) | 1 | (5.3) | 0.23 |
| Cathepsin G | 9 | (32.1) | 2 | (22.2) | 7 | (36.8) | 0.67 |
| VEGF | 23 | (82.1) | 5 | (55.6) | 18 | (94.7) | **0.026** |

Statistical analyses were performed using Fisher's exact test. \*Cases with viable leukemia cells in the brain (CNS involvement). †Cases with viable leukemia cells in CNS or other organs.

We also examined the relationship between the causes of ICH and pathological findings (Table 4). Vascular injury was significantly more frequent in ICH cases caused by leukemia cell infiltration than in those caused by bleeding tendency (75% vs. 0%, p = 0.0005; Table 4). However, no significant differences were observed in the frequency of abnormal permeability or the expression of hemostatic factors in leukemia cells between ICH cases caused by bleeding tendency and those caused by CNS involvement (Table 4).

**Table 4.**
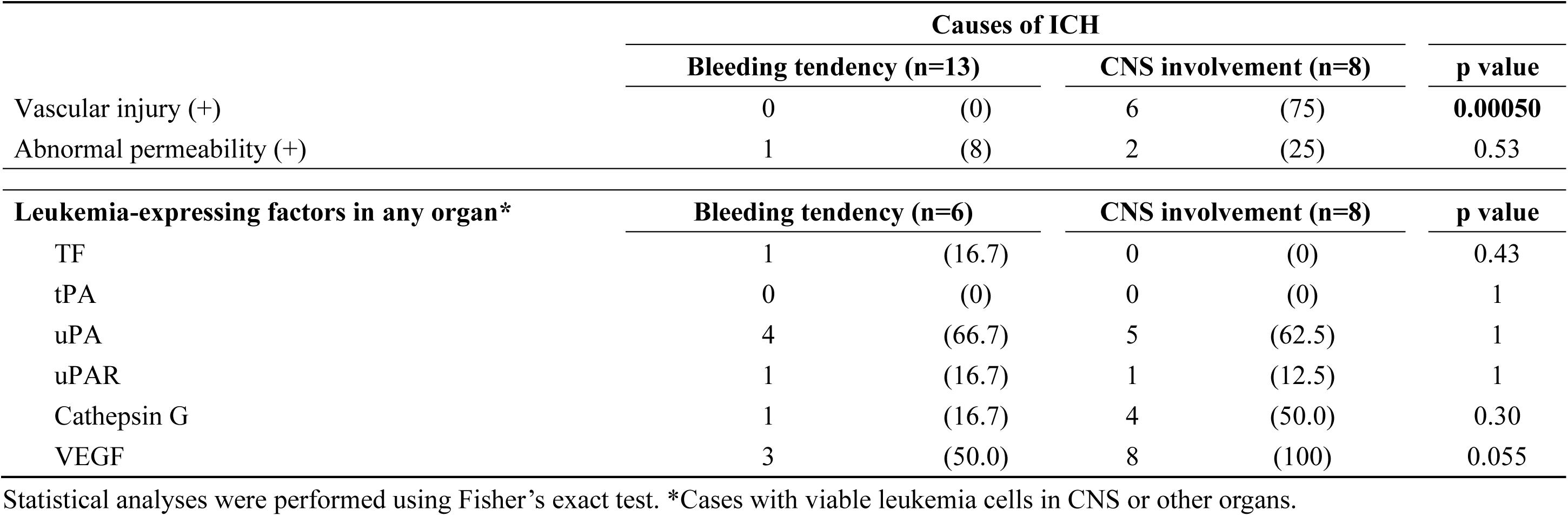
Pathological findings between causes of intracranial hemorrhage (ICH), n (%)

## Discussion

In this autopsy-based study, ICH occurred in 66% of leukemia cases, and CNS involvement was identified in 59%. Among the ICH cases, the underlying cause was attributed to systemic bleeding tendency in 52%, and to direct infiltration of the CNS by leukemia cells in 32%. Notably, 40% of patients with ICH developed fatal brain herniation. A histopathological examination revealed that the leukemia cell infiltration of meninges and vascular walls was associated with direct injury to cerebral vessels, and that vascular injury was frequently associated with leukemic infiltration-related ICH. Furthermore, leukemia cells frequently expressed VEGF, cathepsin G, and uPA. VEGF was highly expressed across all leukemia subtypes and was strongly associated with meningeal invasion. In contrast, cathepsin G expression was primarily observed in myeloid leukemia and associated with vascular injury.

ICH is a major cause of mortality in leukemia.^3^ In the present study, ICH was observed in 66% of leukemia cases, and fatal brain herniation occurred in 40% of those with ICH. These results are consistent with previous findings showing the clinical severity of ICH in hematological malignancies.^1,3,5^ Moreover, ICH in leukemia has been reported to involve various intracranial regions, such as the brain parenchyma, subarachnoid space, and subdural and epidural regions, as well as multifocal sites.^1,24^ A clinical study demonstrated that multifocal cerebral hemorrhage in hematological malignancies was associated with a poor prognosis, and that the mortality rate was high among patients with subarachnoid hemorrhage.^1^ The present study also identified a high frequency of multiple-site ICH and subarachnoid hemorrhage, which is consistent with these findings.^1,24^ In support of a potential mechanistic link, a retrospective cohort study reported a significantly higher incidence of ICH in patients with than in those without CNS involvement (46.1% vs. 3%, p < 0.001).^25^ To further examine this relationship, we pathologically confirmed a high frequency of CNS infiltration by leukemia cells, characterized by the expression of hemostasis-related factors. Moreover, the leukemia cell infiltration of vascular walls was accompanied by the loss or degeneration of medial smooth muscle cells and adventitial collagen. These results suggest that the leukemia cell infiltration of vascular walls makes them vulnerable. These pathological features were present regardless of the ICH status, suggesting that CNS involvement contributes to local vascular injury and increases the risk of ICH even in the absence of clinically evident ICH.

Autopsy-based studies on CNS lesions in leukemia were published in the 1960s and reported that blast crisis and hyperleukocytosis were associated with multiple cerebral hemorrhages and leukemia cell infiltration in the CNS.^12,13,15^ They also identified two characteristic patterns of leukemic involvement: leukostasis and leukemic nodules. In parallel with these early findings, the present study demonstrated that all cases with leukostasis or leukemic nodules were accompanied by ICH; however, the frequency of leukostasis and leukemic nodules did not significantly differ between the ICH and non-ICH groups. This may be due to the small number of cases examined in our study. Kawanami et al. classified the causes of ICH as fungal embolism, leukemic infiltration, and coagulopathy based on the clinical course and autopsy findings.^17^ Our analysis consistently identified systemic bleeding tendency, CNS involvement, fungal embolization, and NBTE as contributing factors. However, previous studies lacked a pathological evaluation of hemorrhagic sites, vascular structures, and CNS involvement, particularly in comparisons with non-ICH or control leukemia cases. To the best of our knowledge, this is the first study to comprehensively evaluate these features in autopsy-confirmed leukemia and identify cerebral vessel injury by leukemia cells and abnormal permeability as novel pathological findings in leukemia cases. The present results suggest that vascular injury, but not abnormal permeability, is associated with the pathogenesis of ICH related to the CNS involvement of leukemia cells.

The CNS involvement of leukemia has been reported in 30–80% of pediatric ALL^26^, 30–40% of adult ALL^27^, and 1.1–30% of AML cases.^28,29^ It is also associated with a poor prognosis.^26–28^ An autopsy study reported CNS involvement in 82% of ALL and 40% of AML cases.^16^ In the present study, CNS involvement was observed in approximately 60% of leukemia cases in a non-remission state at the time of death, and only 5% of these cases had a clinical diagnosis of CNS involvement. Therefore, CNS involvement may occur asymptomatically and potentially cause cerebral vascular injury.

Meningeal invasion is a major route of CNS involvement in leukemia. In a murine model, Yao et al. demonstrated that leukemia cells infiltrated the subarachnoid space by migrating along blood vessels connecting the bone marrow of the skull and spine.^30^ Human autopsy studies also described leukemic localization within the meninges.^13,15,16^ Moore et al. reported subarachnoid and perivascular infiltration in 30 and 19% of autopsy cases with acute leukemia, respectively.^13^ The present study revealed that 50% of leukemia cases exhibited meningeal invasion, specifically infiltration of the subarachnoid space extending to the Virchow–Robin space. However, direct vascular injury caused by meningeal leukemia cells has yet to be reported. In addition to these findings, the present study suggests that intravascular leukemia cells contribute to vascular damage and increased permeability from the luminal side. This is the first study to suggest the leukemia cell infiltration of cerebral vessels from both the outside (via the meningeal route) and inside (via the bloodstream) due to the unique anatomical structure of the brain.

VEGF expression has been reported in various leukemia cell lines.^31^ We herein found that VEGF was frequently expressed by leukemia cells in the brain across various histological types. VEGF plays a critical role in endothelial function and promotes vascular permeability, potentially leading to hemorrhage.^31,32^ Cheng et al. reported that VEGF-overexpressing glioblastoma cells induced ICH in a murine model.^19^ However, VEGF expression did not significantly differ in leukemia cells with or without abnormal permeability or vascular injury in this study. Tumor-derived VEGF also contributes to tumor growth and metastasis.^31,32^ Moreover, high VEGF expression in leukemia cells was associated with meningeal invasion in an ALL murine model.^33^ In that study, the transendothelial migration of leukemia cells was regulated by VEGF and contributed to CNS involvement.^33^ The present study also revealed that VEGF expression in leukemia cells was more frequently observed in cases with than in those without meningeal invasion. Therefore, VEGF appears to affect the leukemia cell infiltration of meninges rather than vascular permeability and injury in humans.

Cathepsin G is a serine protease predominantly found in neutrophils and is involved in host defenses, the regulation of inflammation, vascular homeostasis, and thrombosis.^34^ It also contributes to tissue remodeling by degrading extracellular matrix proteins and activating matrix metalloproteinases.^34^ Shamamian et al. demonstrated that neutrophil-derived cathepsin G activated pro-matrix metalloproteinase-2 and induced microvascular damage *in vitro*.^35^ Furthermore, cathepsin G has been shown to affect the morphology of endothelial cells, inducing intercellular junctional disruption that increases vascular permeability.^35^ Cathepsin G is expressed in leukemia cells, particularly in AML.^20^ The present study demonstrated that cathepsin G was expressed in myeloid leukemia cells and vascular wall-infiltrating cells, and was associated with vascular injury. uPA, a molecule involved in fibrinolysis and extracellular matrix degradation^36^, was also frequently expressed by leukemia cells in the present study; however, it was not associated with vascular injury, abnormal permeability, CNS involvement, or meningeal invasion and, thus, its pathogenetic significance remains unclear. This is the first study based on human autopsy specimens to suggest that leukemia-expressed cathepsin G directly contributed to vascular injury within the brain.

This study has several limitations. Due to its retrospective design and reliance on autopsy specimens, laboratory data at the time of death were not consistently available, which, along with the small sample size, limited the ability to detect significant relationships and correlate pathological findings with hemostasis-related parameters. Furthermore, leukemia subtypes were classified using clinical and histological data, without the full immunophenotypic or genetic information needed for classification under the 2022 WHO criteria. Future studies integrating standardized laboratory and molecular data may provide further insights into the relationship between the CNS pathology and leukemia subtypes. Additionally, the small number of acute promyelocytic leukemia cases, a subtype with a high hemorrhagic risk, may limit the generalizability of our results to this population.

In conclusion, this autopsy-based study revealed CNS involvement in a large percentage of leukemia cases that was frequently accompanied by ICH. Leukemia cell infiltration, via both meningeal and intravascular routes, was pathologically associated with cerebral vascular injury. Cerebral vascular injury may represent a mechanism of ICH associated with CNS involvement by leukemia cells. Importantly, we identified VEGF and cathepsin G as potential markers of meningeal invasion and cerebral vascular damage in leukemia, respectively. These results provide novel insights into the mechanisms underlying CNS vascular pathology and hemorrhagic complications in leukemia.

## Data Availability

All data produced in the present study are available upon reasonable request to the authors

## Acknowledgments

We are grateful to Nahoko Udatsu, Kyoko Ohashi, and Hayato Ijichi for their technical assistance.

This study was partly supported by Grants-in-Aid for Scientists from the Japan Society for the Promotion of Science (JSPS KAKENHI Grant Numbers 18K15083, 19K07437, 20K08085, 21K15403, 21K07706, 23K06467), and a Clinical Research Support Grant from University of Miyazaki Hospital.

## Authorship

T. G, Y. H., and A. Y. contributed to the conception and design of the study; T. G, K. K., K. S., E. N., N. O., M. A., K. M., S. MG, and K. S. participated in data collection and analysis; T. G. wrote the first draft of the manuscript; Y. H. and A. Y. reviewed the manuscript critically; and all authors read and approved the final manuscript.

Conflict-of-interest disclosure: The authors declare no competing financial interests.

## Supplemental Material

Tables S1-S5

Figure S1

