## Supplemental Figures for "Cathepsin G is associated with cerebral vascular injury in leukemia: A pathological insight into intracranial hemorrhage"

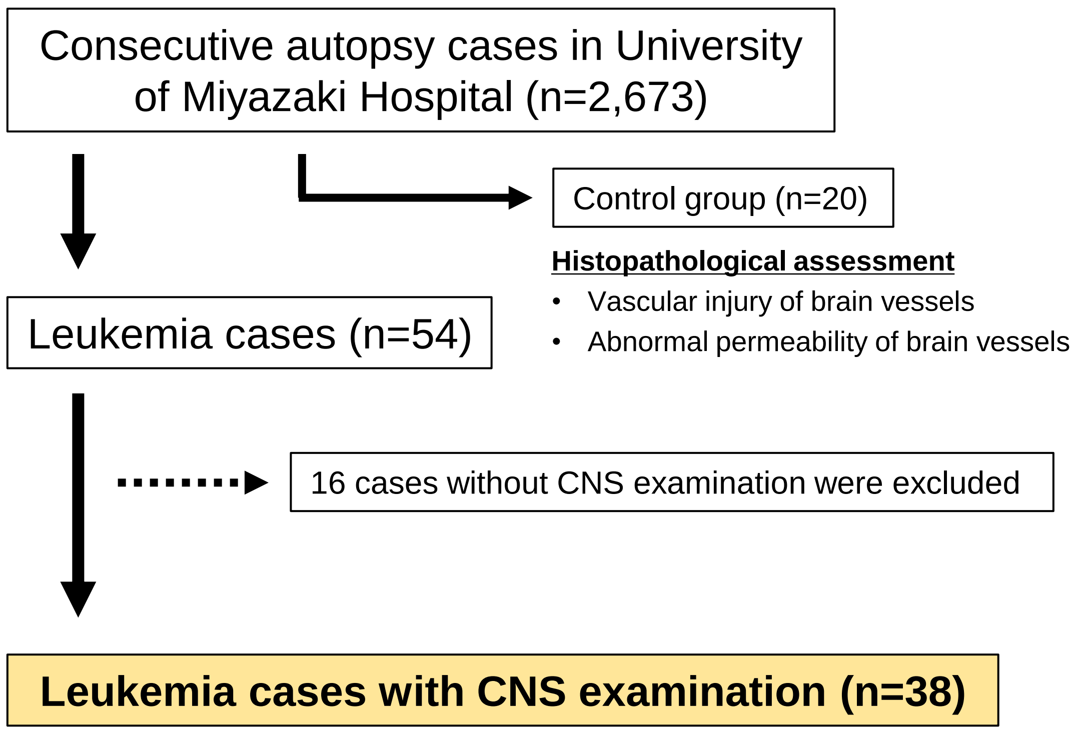


**Analysis of autopsy-record summary (n=38)**

- Clinical history
- Leukemia classification
- Presence/localization of hemorrhage in CNS

**Histopathological assessment**

- Presence/localization of leukemia cell in CNS (n=38)
- Vascular injury of brain vessels (n=38)
  - Smooth muscle actin (IHC) and Sirius-red stain (for collagen fiber)
- Abnormal permeability of brain vessels (n=38)
  - Perivascular deposition of fibrin or von-Willebrand factor (IHC)
- Expression of hemostatic/ vascular factors and protease in leukemia cells (n=28)
  - VEGF, cathepsin G, tPA, uPA, uPAR, tissue factor (IHC)
- Double immunofluorescence (representative cases)

**Figure S1. Summary of study design**
