## Supplemental Tables for "Cathepsin G is associated with cerebral vascular injury in leukemia: A pathological insight into intracranial hemorrhage"

**Table S1. Primary antibodies for immunohistochemistry and immunofluorescence**

| Antibody | Antigen/Marker | Species | Clone | Dilution | HIER | Company | Catalog number |
|---|---|---|---|---|---|---|---|
| VEGF (A-20) | vascular endothelial growth factor | rabbit | polyclonal | 1:500 | MW | Santa Cruz Biotechnology | Sc-152 |
| Cathepsin G | a neutrophil serine protease | rabbit | E3N30 | 1:500 | MW | Cell Signaling Technology | P08311 |
| tPA | tissue plasminogen activator | rabbit | polyclonal | 1:100 | MW | GeneTex | GTX103453 |
| uPA | urokinase-type plasminogen activator | rabbit | polyclonal | 1:50 | MW | Atlas Antibodies | HPA008719 |
| uPAR | urokinase-type plasminogen activator receptor | mouse | E-3 | 1:50 | MW | Santa Cruz Biotechnology | Sc-376494 |
| Tissue factor | tissue factor | mouse | H-9 | 1:200 | MW | Santa Cruz Biotechnology | Sc-374441 |
| SMA | α-smooth muscle actin, the median of vessels | mouse | 1A4 | 1:200 | (-) | Agilent/Dako | M0851 |
| VWF | von Willebrand factor | mouse | 36B11 | 1:100 | MW | Leica Biosystems | NCL-L-vWF |
| Fibrin | fibrin | mouse | 59D8 | 1:1000 | MW | EMD Millipore Corp. | MABS2155-25-UG |
| Myeloperoxidase | myeloperoxidase | rabbit | polyclonal | 1:1000 | (-) | Agilent/Dako | A0398 |
| MPO light chain | myeloperoxidase | mouse | A-5 | 1:200 | (+) | Santa Cruz Biotechnology | Sc-365436 |

HIER, heat-induced epitope retrieval methods; MW, microwave. MPO light chain is only used for immunofluorescence.

**Table S2.**
**Clinicopathological background of the control group (n=20)**

| **Clinical background** | | |
|---|---|---|
| Age, y, median (range) | 64.0 | (18-76) |
| Male, n (%) | 15 | (75.0) |
| Cancer-bearing status, n (%) | 9 | (45.0) |
| Primary organ of cancer, n (%) | | |
| *Lung* | *3* | *(15.0)* |
| *Gastrointestinal system* | *3* | *(15.0)* |
| *Thyroid gland* | *2* | *(10.0)* |
| *Skin* | *1* | *(5.0)* |
| Chemotherapy, n (%) | 9 | (45.0) |
| Interstitial pneumonia, n (%) | 3 | (15.0) |
| Systemic vasculitis, n (%) | 1 | (5.0) |
| Severe infection, n (%) | 7 | (35.0) |
| Systemic hemorrhage, n (%) | 7 | (35.0) |
| Multiple microthrombi, n (%) | 0 | (0) |
| DVT, n (%) | *3* | (15.0) |
| PE, n (%) | 4 | (20.0) |
| NBTE, n (%) | 0 | (0) |

DVT, deep vein thrombosis; PE, pulmonary embolism; NBTE, non-bacterial thrombotic endocarditis.

*Severe infection was defined as an infectious disease diagnosed clinically and pathologically, which was related to the cause of death.

†Multiple microthrombi was defined as the presence of microthrombi in more than three organs.

**Table S3. Comparative analysis of clinicopathological characteristics between control and leukemia groups**

| | Control group (n=20) | | Leukemia group (n=38) | | P value |
|---|---|---|---|---|---|
| Age, y, median (range) | 64 | (18-76) | 58 | (13-82) | 0.26 |
| Male, n (%) | 15 | (75.0) | 29 | (76.3) | 0.75 |
| Chemotherapy, n (%) | 9 | (45.0) | 33 | (86.8) | 0.0015 |
| Severe infection, n (%) | 7 | (35.0) | 24 | (63.2) | 0.055 |
| Systemic hemorrhage, n (%) | 7 | (35.0) | 20 | (52.6) | 0.27 |
| Multiple microthrombi, n (%) | 0 | (0) | 5 | (13.2) | 0.15 |
| DVT, n (%) | 3 | (15.0) | 0 | (0) | 0.04 |
| PE, n (%) | 4 | (20.0) | 4 | (10.5) | 0.43 |
| NBTE, n (%) | 0 | (0) | 2 | (5.3) | 0.54 |

DVT, deep vein thrombosis; PE, pulmonary embolism; NBTE, non-bacterial thrombotic endocarditis.

Statistical analyses were performed using Fisher's exact test.

**Table S4. Clinicopathological findings of leukemia cases with intracranial hemorrhage (n=25)**

| | | |
|---|---|---|
| **Leukemia type, n (%)** | | |
| AML | 16 | (64.0) |
| ALL | 5 | (20.0) |
| CML, blast phase | 4 | (16.0) |
| **Leukemia status, n (%)** | | |
| Non-remission state of leukemia | 23 | (92.0) |
| *CNS involvement of leukemia cells* | *11* | *(44.0)* |
| **Sites of intracranial hemorrhage*, n (%)** | | |
| Brain parenchyma | 19 | (76.0) |
| Subarachnoid extending to the Virchow-Robin space | 16 | (64.0) |
| Intraventricular space | 7 | (28.0) |
| Subdural | 3 | (12.0) |
| Multiple sites | 15 | (60.0) |
| Complicated with fetal brain herniation | 10 | (40.0) |
| **Causes of intracranial hemorrhage, n (%)** | | |
| Bleeding tendency | 13 | (52.0) |
| CNS involvement of leukemia | 8 | (32.0) |
| Aspergillosis | 2 | (8.0) |
| Emboli from NBTE (hemorrhagic infarction) | 1 | (4.0) |
| Mucormycosis + CNS involvement of leukemia | 1 | (4.0) |

*Cases involved multiple sites.

**Table S5. Immunohistochemical expression of hemostatic factors in leukemia cells, n (%)**

| | | VEGF | | Cathepsin G | | tPA | | uPA | | uPAR | | TF | |
|---|---|---|---|---|---|---|---|---|---|---|---|---|---|
| Total | n=28 | 23 | (82.1) | 9 | (32.1) | 1 | (3.6) | 22 | (78.6) | 3 | (10.7) | 2 | (7.1) |
| Leukemia type | | | | | | | | | | | | | |
| AML | n=17 | 16 | (94.1) | 7 | (41.2) | 1 | (5.9) | 14 | (82.4) | 1 | (5.9) | 2 | (11.8) |
| ALL | n=6 | 4 | (66.7) | 0 | (0.0) | 0 | (0.0) | 4 | (66.7) | 0 | (0.0) | 0 | (0.0) |
| CML, blast phase | n=4 | 3 | (75.0) | 2 | (50.0) | 0 | (0.0) | 3 | (75.0) | 2 | (50.0) | 0 | (0.0) |
| CLL/SLL | n=1 | 0 | (0.0) | 0 | (0.0) | 0 | (0.0) | 1 | (100.0) | 0 | (0.0) | 0 | (0.0) |

**Table S6. Relationships among intracranial hemorrhage (ICH), pathological findings, and the expression of hemostatic factors, n (%)**

| **Pathological findings** | **Total (n=38)** | | **Non-ICH (n=13)** | | **ICH (n=25)** | | **p value** |
|---|---|---|---|---|---|---|---|
| Non-remission state | 34 | (89.5) | 11 | (84.6) | 23 | (92.0) | 0.59 |
| CNS involvement | 20 | (52.6) | 8 | (61.5) | 12 | (48.0) | 0.51 |
| Vascular injury | 13 | (34.2) | 5 | (38.5) | 8 | (32.0) | 0.73 |
| Abnormal permeability | 6 | (15.8) | 1 | (7.7) | 5 | (20.0) | 0.64 |
| **Leukemia-expressing factors in any organ*** | **Total (n=28)** | | **Non-ICH (n=11)** | | **ICH (n=17)** | | **p value** |
| TF | 2 | (7.1) | 1 | (9.1) | 1 | (5.9) | 1 |
| tPA | 1 | (3.6) | 1 | (9.1) | 0 | (0.0) | 0.39 |
| uPA | 22 | (78.6) | 11 | (100.0) | 11 | (64.7) | 0.055 |
| uPAR | 3 | (10.7) | 1 | (9.1) | 2 | (11.8) | 1 |
| Cathepsin G | 9 | (32.1) | 3 | (27.3) | 6 | (35.3) | 1 |
| VEGF | 23 | (82.1) | 9 | (81.8) | 14 | (82.4) | 1 |
| **Leukemia-expressing factors in the brain†** | **Total (n=20)** | | **Non-ICH (n=8)** | | **ICH (n=12)** | | **p value** |
| TF | 0 | (0.0) | 0 | (0.0) | 0 | (0.0) | 1 |
| tPA | 1 | (5.0) | 1 | (12.5) | 0 | (0.0) | 0.4 |
| uPA | 16 | (80.0) | 8 | (100.0) | 8 | (66.7) | 0.12 |
| uPAR | 1 | (5.0) | 0 | (0.0) | 1 | (8.3) | 1 |
| Cathepsin G | 7 | (35.0) | 2 | (25.0) | 5 | (41.7) | 0.64 |
| VEGF | 18 | (90.0) | 7 | (87.5) | 11 | (91.7) | 1 |
| **Leukemia involvement in the brain†** | **Total (n=20)** | | **Non-ICH (n=8)** | | **ICH (n=12)** | | **p value** |
| Leukostasis | 4 | (20.0) | 0 | (0.0) | 4 | (33.3) | 0.12 |
| Leukemic nodule | 4 | (20.0) | 0 | (0.0) | 4 | (33.3) | 0.12 |

Statistical analyses were performed using Fisher's exact test. *Cases with viable leukemia cells in CNS or other organs.

†Cases with viable leukemia cells in the brain (CNS involvement).
